# Acceptability of the routine use of pulse oximeters in the Integrated Management of Childhood Illness guidelines at primary health centers in West Africa: a mixed-methods study

**DOI:** 10.1101/2024.10.16.24315522

**Authors:** Sarah Louart, Gildas Boris Hedible, Habibata Balde, Abdourahmane Coulibaly, Abdoua Elhadji Dagobi, Kadidiatou Kadio, Désiré Neboua, Zineb Zair, Valériane Leroy, Valéry Ridde, AIRE study group

## Abstract

**Introduction:** To better identify illnesses with severe hypoxemia in children aged under-5 years, the AIRE project has implemented the routine use of a Pulse Oximeter (PO) into the Integrated Management of Childhood Illness (IMCI) guidelines at primary health centers (PHC) in Burkina Faso, Guinea, Mali, and Niger. We aimed to measure and understand the acceptability among healthcare workers (HCW) and children’s families (CF) of using PO in these contexts.

**Methods:** Based on an original conceptual framework, we conducted a convergent mixed methods study to assess HCW’s and CF’s acceptability. We used quantitative questionnaires based on the Likert scale to conduct four repeated cross-sectional studies among all HCW on duty in the 202 PHC involved in the AIRE project before the intervention (before and after PO use training), six months after the introduction of the PO, and two months after project completion. We also conducted semi-structured interviews with HCW (n=102) and CF (n=59).

**Results:** From March 2021 to December 2022, 486, 537, 538 and 476 HCW completed the four acceptability surveys. Overall, 31% of HCW had mitigated feelings about PO use before the training, 46% found it rather acceptable and 23% strongly acceptable. At the end of the project, it was respectively 15%, 34% and 51%. PO training was consistently associated with greater HCW acceptability. HCW found many advantages in using PO, such as improved care for children, more accurate diagnosis, and a boost in their confidence in childcare management. Nevertheless, perceived increased workload and consultation time, as well as difficulties in referring children to hospital remain challenging. CF did not necessarily understand the new device’s purpose, but their opinions of the technology were generally positive.

**Conclusion:** The PO use, integrated into IMCI consultations, seems to be accepted by HCW and CF, although sustainable challenges remain.

**What is already known on this topic:**

- Even if PO effectiveness is well-known, integration of new medical technologies in primary health care settings in low-resource countries often faces challenges, including acceptability among healthcare workers and families.
- Previous studies that have looked at the acceptability of PO were mostly hospital-based and did not use a comprehensive methodology to investigate acceptability in depth.

**What this study adds:**

- To our knowledge, this is the first study to examine the acceptability of a healthcare innovation to HCW and CF, based on a comprehensive theoretical framework, combining a quantitative and qualitative approach, and using an innovative acceptability score.
- This study provides empirical evidence on the high acceptability of PO use for HCW and CF at a decentralized level in Burkina Faso, Guinea, Mali, and Niger.
- Training and supervision significantly improved HCW acceptability of PO use, with increased confidence in childcare management. However, challenges such as increased workload and consultation time, as well as referral difficulties, persist despite the overall positive opinion.

**How this study might affect research, practice or policy:**

- The findings support the need for comprehensive training and supervision programs to enhance acceptability among HCW.
- Policymakers should work towards addressing the operational challenges identified, especially those related to the referral of children, which could hinder the sustainability of PO use.
- Research aiming to evaluate acceptability should be based on conceptual frameworks or theories that allow for an in-depth exploration of the different dimensions of acceptability.

## Introduction

Acute respiratory infections (ARI) are among the leading causes of death in children under five, mainly in low-middle income countries [1,2]. Technological innovations in healthcare can enhance prevention and treatment capacities, and the overall quality of services [3]. Among these devices, the pulse oximeter (PO) is crucial for measuring oxygen saturation levels and identifying hypoxemia (low oxygen saturation in the blood). Severe hypoxemia, common in ARI and illnesses like malaria and malnutrition, is a life-threatening condition that requires urgent treatment. Therefore, improving access to hypoxemia diagnostic tools (oximetry) and treatment (oxygen therapy), has long been considered crucial [4]. However, many studies highlight significant gaps in oximetry and oxygen access in Africa [5–9]. While PO are used daily in high-resource countries, their use in frontline facilities remains relatively unexplored in West Africa.

The AIRE project (2019-2022) implemented the routine use of PO into the Integrated Management of Childhood Illness (IMCI) algorithms in 202 primary health centers (PHC) and eight hospitals in Burkina Faso, Guinea, Mali and Niger [10]. The PO model used in this project was Acare Technology (Taiwan; AH-M1 S0002033) that works with probes. The successful introduction of new devices is closely linked to their acceptability, which is a multidimensional concept [11–13] crucial for understanding the processes involved in healthcare innovations dissemination [14]. Acceptability analysis must go beyond mere satisfaction with the technology; it includes factors like perceived ease of use and compatibility with local health routines. It is influenced by individual experiences, societal norms, the healthcare system’s infrastructure, etc. [15].

Few studies have explored the acceptability of PO in LMIC. Existing research is often concentrated at the hospital level rather than decentralized [8,16,17]. Some emerging studies explore the barriers and facilitators of PO use among community health workers and PHC [18–20]. However, to our knowledge, no studies are reported at the PHC level in West Africa. Furthermore, not all studies directly address acceptability, and none use a solid conceptual framework. This gap highlights the need for more research to understand the broader applicability and challenges of implementing PO in these settings. Our study aimed to evaluate the acceptability of PO use among healthcare workers (HCW) and children’s families (CF), identifying both barriers and facilitators to its adoption.

## Methods

This study was based on a conceptual framework and utilized mixed methods to ensure completeness and enhance explanations [21]. We employed a convergent design and the Mixed Methods Appraisal Tool to ensure accurate reporting of the various elements [22].

### Conceptual framework

We used a conceptual framework that identifies six dimensions of the acceptability of technological healthcare innovations [23]: compatibility, perceived advantages, personal emotions, social influence, perceived disadvantages, and perceived complexity, all influenced by the context and the form of the intervention. For this study, we also adopted the three temporal perspectives for acceptability proposed by Sekhon et al. [15]: prospective (assessed before implementation, focusing on initial perceptions and willingness to engage), concurrent (during use, capturing real-time experiences and attitudes) and retrospective acceptability (after completion of intervention, reflecting on overall experience and satisfaction).

### Study sites

The study was conducted in two health districts per country [10], in the 202 PHC. All children attending IMCI consultations, except those aged 2 to 59 months classified as IMCI green cases without respiratory symptoms, were eligible for PO use. Those with severe hypoxemia (defined by SpO2<90%) should be transferred to hospital for urgent oxygen therapy. The size of PHC was determined by the number of consultations for children under-5 in 2019, classified as small (≤1,000 visits), medium (1,001-3,000 visits), or large (>3,000 visits) [24]. The quantitative study on the acceptability of PO use covered all PHC and their referral district hospital (n=8). Qualitative data collection involved all district hospitals and 16 PHC selected as research sites (four per country) [10].

### Study design and inclusion process

We conducted repeated cross-sectional individual surveys for quantitative data from March 2021 to December 2022. For the prospective acceptability survey, all PHC managers, their deputies, and HCW involved in IMCI consultations received classroom training on PO use and an IMCI practice update over 6 days to 2 weeks, depending on the country. Data were collected before and after training. The other two surveys were conducted six months after the start of the project (concurrent acceptability) and two months after the project’s end (retrospective acceptability). Surveys included all the HCW in the PHC at the time of the survey. From June 2021 to December 2022, we also measured the proportion of PO use among eligible children using monthly aggregated data from IMCI consultation registers (electronic or paper-based) in all PHC.

The qualitative study was conducted in two phases. The concurrent acceptability period (T1) involved semi-structured interviews with HCW and CF seen in consultations and the retrospective acceptability period (T2) included interviews only with HCW to understand how their acceptability changes over time. Observations of PO use during consultations were also conducted to assess HCW familiarity and proficiency, interactions with CF, and children’s reactions to PO. All HCW in the research sites conducting consultations with children under five and available at the time of the survey were interviewed. To select CF, we used purposive sampling [25] to ensure representativeness according to variables such as sex, age, relationship with the child, distance to the PHC and the child’s health status. Empirical saturation determined the final number of participants [26].

### Data collection

Quantitative data were collected using REDCap software, focusing on HCW’s socio-demographic characteristics, prior IMCI practices, knowledge and previous use of PO, and specific data to assess PO acceptability. Aggregated data included the number of children eligible for PO use and the frequency of its use by HCW.

National co-investigators conducted qualitative interviews in local languages or French, fully transcribed and translated when necessary.

### Calculation of an acceptability score

Quantitative questions for five of the dimensions [23] (ranging from 2 to 5), except for the “social influence” dimension, were combined to construct each acceptability dimension (Appendix 1). They were based on a five-modalities Likert scale [27]. Each option was assigned a score from −10 to 10. By compiling and averaging the scores for questions in each dimension, we obtained a score for each acceptability dimension. Composite reliability was assessed to evaluate the consistency of questions included within each dimension [28] (Appendix 2). Dimensions scores were then aggregated using the same logic and averaged to derive the overall acceptability score, categorized according to the Likert scale as “strongly unacceptable”, “unacceptable “, “mixed feelings”, “acceptable “ and “strongly acceptable”. Data collection on the social influence dimension was not entirely conducted using the Likert scale format and proved challenging to measure prospectively. Therefore, we will only describe the various possible influences observed.

### Data analysis

We described and compared by country the socio-demographic characteristics of the HCW enrolled in the concurrent study, the most representative of the four surveys. We described the dimensions of acceptability and the overall score’s evolution over time and by country. Quantitative variables were described using means and standard deviations. Categorical data were presented as proportions with their 95% confidence intervals and compared using Pearson’s χ2 or Fisher’s exact tests. Using an ordinal logistic regression model with a random effect for the health district (only for the model combining data from the four countries), we explored the factors associated with “strongly acceptable” versus lower acceptability levels. The main variables included in the modelling were age, sex, profession, years of experience, type of IMCI support (electronic vs. paper-based), prior knowledge of PO, previous training on PO use, and the size of PHC. Sex and age were forced in all models. Analysis was performed using R software version 4.3.0. and considered statistically significant with a p-value<0.05.

All qualitative data were analyzed using Nvivo12 software. The research team developed a common codebook and conducted a thematic analysis to compare the empirical data with the conceptual framework [29]. Coding was initially done by country, followed by a cross-country comparative analysis.

We then analyzed acceptability by integrating qualitative and quantitative results across the various dimensions of acceptability and its associated factors. This approach allowed for comparison of data, analyzing convergence, divergence and explanation, facilitating a richer and more comprehensive interpretation [22].

### Ethical aspects

This study was approved by the national ethics committees of the four countries, along with the WHO and INSERM committees [10]. It was also registered in the Pan African Clinical Trial Registry. Participants were included with their free and informed written consent.

### Patient and public involvement

Individual data were collected with authorization from the ethics committees and ministries of health. Patients were not involved in the analysis, interpretation of results, or writing of the manuscript.

## Results

The quantitative surveys included 486 (pre-training), 537 (post-training), 538 (concurrent) and 476 (retrospective) HCW over time. Interviews were conducted with 50 HCW and 59 CF (concurrent), then 52 HCW (retrospective) (Figure 1).

**Figure 1:**
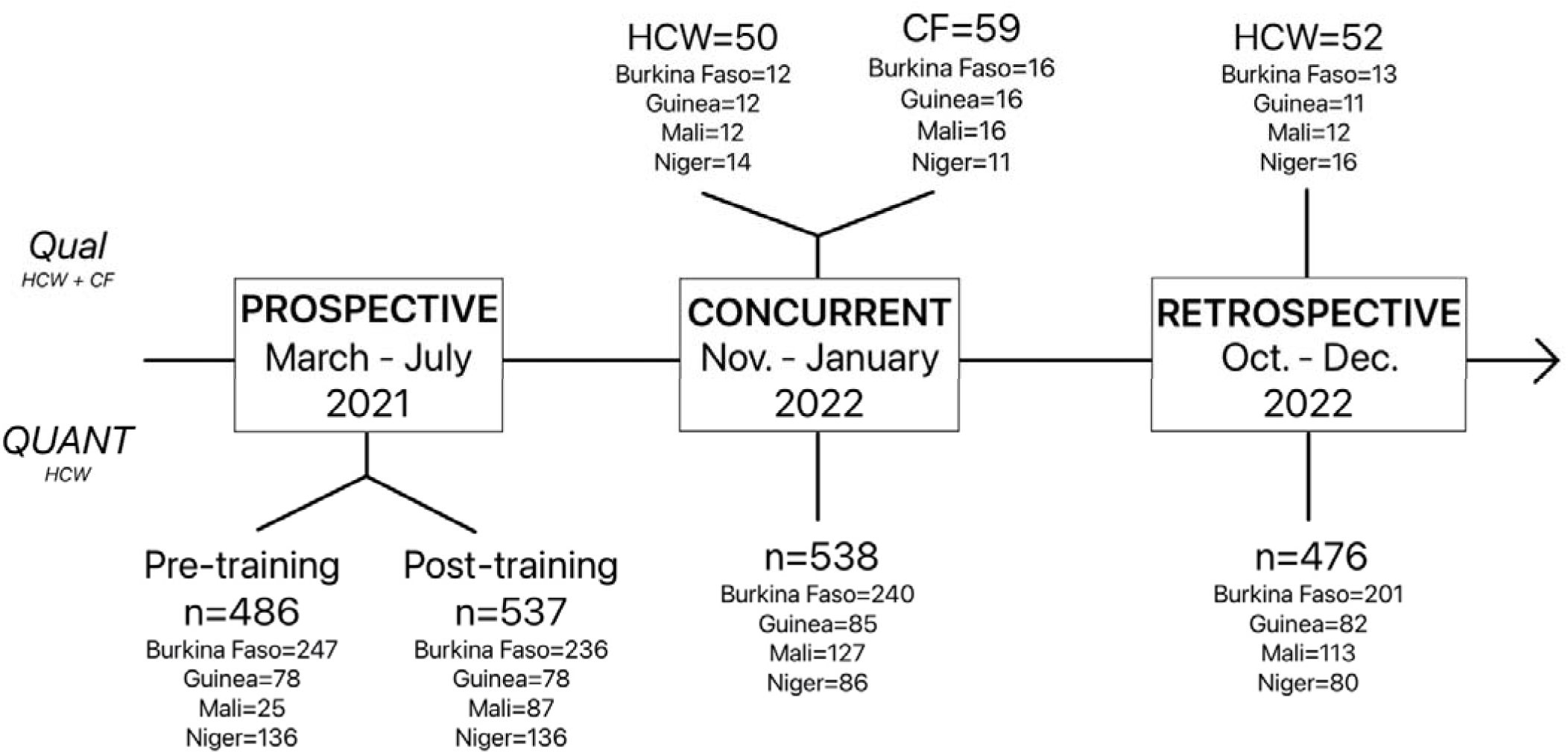
Timeline of data collection phases and number of participants.

### HCW socio-demographic characteristics

Out of the 538 HCWs enrolled in the concurrent acceptability survey, 53.2% were aged 25-35 years, ranging from 41.9% in Niger to 61.2% in Guinea. Women represented 47.8% overall, with significant country variations (Table 1). All worked in rural settings, except for Guinea and Niger, where 52.9% and 25.6% of HCW, respectively, were from urban areas. Overall, 44.8% of HCW had one to five years of experience. Medical doctors and nurses accounted for 63.0% of all HCW. Before the initial PO training, 75.5% of HCW were unaware of the PO. Six months after the intervention started, 77.1% had been trained overall.

**Table 1.**
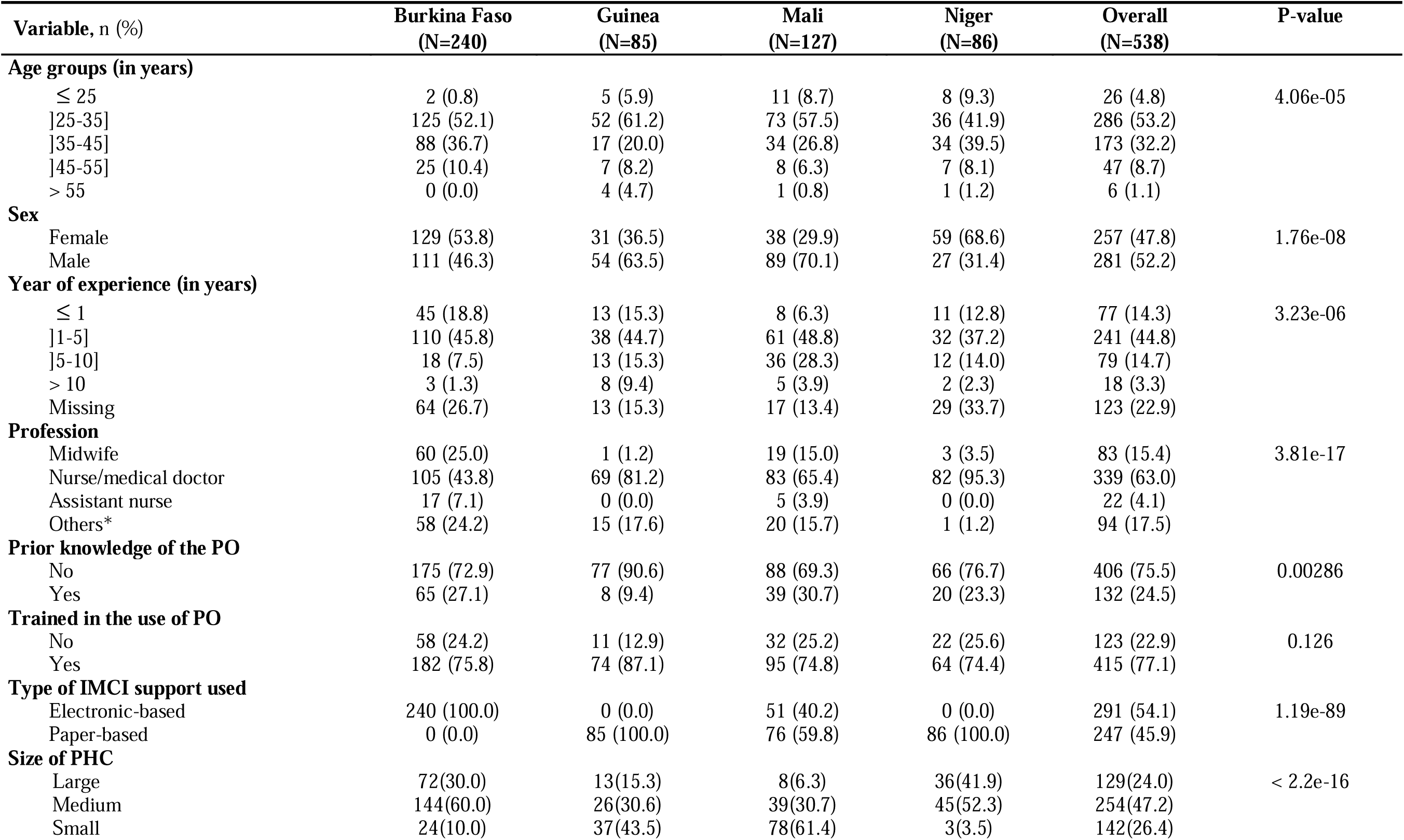
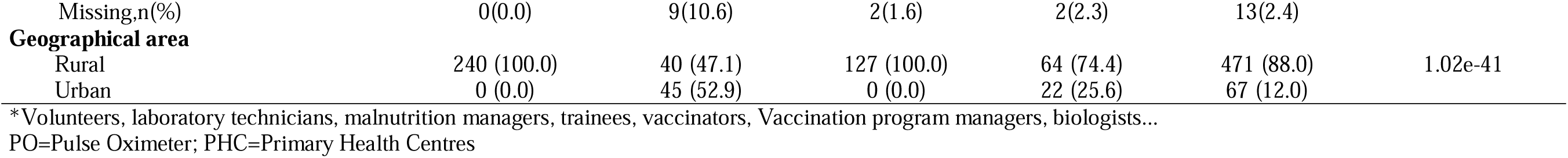
Socio-demographic characteristics of Health Care Workers during the concurrent acceptability of Pulse Oximeters use survey (N=538), conducted in the 202 AIRE Primary Health Centres, November 2021-January 2022.

| Variable, n (%) | Burkina Faso<br>(N=240) | Guinea<br>(N=85) | Mali<br>(N=127) | Niger<br>(N=86) | Overall<br>(N=538) | P-value |
| --- | --- | --- | --- | --- | --- | --- |
| **Age groups (in years)** |  |  |  |  |  |  |
| ≤ 25 | 2 (0.8) | 5 (5.9) | 11 (8.7) | 8 (9.3) | 26 (4.8) | 4.06e-05 |
| ]25-35] | 125 (52.1) | 52 (61.2) | 73 (57.5) | 36 (41.9) | 286 (53.2) |  |
| ]35-45] | 88 (36.7) | 17 (20.0) | 34 (26.8) | 34 (39.5) | 173 (32.2) |  |
| ]45-55] | 25 (10.4) | 7 (8.2) | 8 (6.3) | 7 (8.1) | 47 (8.7) |  |
| > 55 | 0 (0.0) | 4 (4.7) | 1 (0.8) | 1 (1.2) | 6 (1.1) |  |
| **Sex** |  |  |  |  |  |  |
| Female | 129 (53.8) | 31 (36.5) | 38 (29.9) | 59 (68.6) | 257 (47.8) | 1.76e-08 |
| Male | 111 (46.3) | 54 (63.5) | 89 (70.1) | 27 (31.4) | 281 (52.2) |  |
| **Year of experience (in years)** |  |  |  |  |  |  |
| ≤ 1 | 45 (18.8) | 13 (15.3) | 8 (6.3) | 11 (12.8) | 77 (14.3) | 3.23e-06 |
| ]1-5] | 110 (45.8) | 38 (44.7) | 61 (48.8) | 32 (37.2) | 241 (44.8) |  |
| ]5-10] | 18 (7.5) | 13 (15.3) | 36 (28.3) | 12 (14.0) | 79 (14.7) |  |
| > 10 | 3 (1.3) | 8 (9.4) | 5 (3.9) | 2 (2.3) | 18 (3.3) |  |
| Missing | 64 (26.7) | 13 (15.3) | 17 (13.4) | 29 (33.7) | 123 (22.9) |  |
| **Profession** |  |  |  |  |  |  |
| Midwife | 60 (25.0) | 1 (1.2) | 19 (15.0) | 3 (3.5) | 83 (15.4) | 3.81e-17 |
| Nurse/medical doctor | 105 (43.8) | 69 (81.2) | 83 (65.4) | 82 (95.3) | 339 (63.0) |  |
| Assistant nurse | 17 (7.1) | 0 (0.0) | 5 (3.9) | 0 (0.0) | 22 (4.1) |  |
| Others* | 58 (24.2) | 15 (17.6) | 20 (15.7) | 1 (1.2) | 94 (17.5) |  |
| **Prior knowledge of the PO** |  |  |  |  |  |  |
| No | 175 (72.9) | 77 (90.6) | 88 (69.3) | 66 (76.7) | 406 (75.5) | 0.00286 |
| Yes | 65 (27.1) | 8 (9.4) | 39 (30.7) | 20 (23.3) | 132 (24.5) |  |
| **Trained in the use of PO** |  |  |  |  |  |  |
| No | 58 (24.2) | 11 (12.9) | 32 (25.2) | 22 (25.6) | 123 (22.9) | 0.126 |
| Yes | 182 (75.8) | 74 (87.1) | 95 (74.8) | 64 (74.4) | 415 (77.1) |  |
| **Type of IMCI support used** |  |  |  |  |  |  |
| Electronic-based | 240 (100.0) | 0 (0.0) | 51 (40.2) | 0 (0.0) | 291 (54.1) | 1.19e-89 |
| Paper-based | 0 (0.0) | 85 (100.0) | 76 (59.8) | 86 (100.0) | 247 (45.9) |  |
| **Size of PHC** |  |  |  |  |  |  |
| Large | 72(30.0) | 13(15.3) | 8(6.3) | 36(41.9) | 129(24.0) | < 2.2e-16 |
| Medium | 144(60.0) | 26(30.6) | 39(30.7) | 45(52.3) | 254(47.2) |  |
| Small | 24(10.0) | 37(43.5) | 78(61.4) | 3(3.5) | 142(26.4) |  |
| Missing,n(%) | 0(0.0) | 9(10.6) | 2(1.6) | 2(2.3) | 13(2.4) |  |
| **Geographical area** |  |  |  |  |  |  |
| Rural | 240 (100.0) | 40 (47.1) | 127 (100.0) | 64 (74.4) | 471 (88.0) | 1.02e-41 |
| Urban | 0 (0.0) | 45 (52.9) | 0 (0.0) | 22 (25.6) | 67 (12.0) |  |

\*Volunteers, laboratory technicians, malnutrition managers, trainees, vaccinators, Vaccination program managers, biologists...
PO=Pulse Oximeter; PHC=Primary Health Centres

### PO uptake

Monthly trends of PO use show that it quickly reached and exceeded 80% for eligible children, except in Burkina Faso, where it took around six months to reach this target. Overall, PO use reached 93.4% of the 381,874 eligible children attending PHC over the study period (Appendix 3).

### HCW acceptability

#### Overall acceptability

Overall acceptability scores for the PO were high across data collections (Table 2). No HCW found the PO “not at all acceptable” or “not acceptable”. The proportion of HCW with mixed feelings decreased over time, while those finding the PO “strongly acceptable” increased from 22.6% (pre-training) to 51.3% (retrospective) (p-value<0.001). This positive trend is also reflected in improving “strongly acceptable” rates between the first and last data collections; however, the paths diverge depending on the country. In Burkina Faso, this improvement was rather linear. In contrast, in the other three countries, there was a notable improvement after the training followed by a decline during the concurrent data collection. Finally, the paths diverged, with overall acceptability rising significantly in Guinea (which ultimately had the highest acceptability rate) and slightly in Niger for the final collection while continuing to decrease in Mali.

**Table 2.** Overall acceptability score combining the five dimensions except social influence of Health Care Workers to Pulse Oximeters use according to country and time of data collection in the 202 AIRE Primary Health Centres, March 2021-December 2022.

| Time of data collection, n (%) | Burkina Faso | Guinea | Mali | Niger | Overall | P-value |
| --- | --- | --- | --- | --- | --- | --- |
| **Prospective pre-training (n)** | **247** | **78** | **25** | **136** | **486** |  |
| Strongly unacceptable | 0 (0.0) | 0 (0.0) | 0 (0.0) | 0 (0.0) | 0 (0.0) | 0.523 |
| Unacceptable | 0 (0.0) | 0 (0.0) | 0 (0.0) | 0 (0.0) | 0 (0.0) |  |
| Mixed feelings | 74 (30.0) | 27 (34.6) | 12 (48.0) | 39 (28.7) | 152 (31.3) |  |
| Acceptable | 115 (46.5) | 32 (41.0) | 9 (36.0) | 68 (50.0) | 224 (46.1) |  |
| Strongly acceptable | 58 (23.5) | 19 (24.4) | 4 (16.0) | 29 (21.3) | 110 (22.6) |  |
| **Prospective post-training (n)** | **236** | **78** | **87** | **136** | **537** |  |
| Strongly unacceptable | 0 (0.0) | 0 (0.0) | 0 (0.0) | 0 (0.0) | 0 (0.0) | 0.007 |
| Unacceptable | 0 (0.0) | 0 (0.0) | 0 (0.0) | 0 (0.0) | 0 (0.0) |  |
| Mixed feelings | 42 (17.8) | 13 (16.7) | 18 (20.7) | 14 (10.3) | 87 (16.2) |  |
| Acceptable | 111 (47.0) | 38 (48.7) | 42 (48.3) | 49 (36.0) | 240 (44.7) |  |
| Strongly acceptable | 83 (35.2) | 27 (34.6) | 27 (31.0) | 73 (53.7) | 210 (39.1) |  |
| **Concurrent (n)** | **240** | **85** | **127** | **86** | **538** |  |
| Strongly unacceptable | 0 (0.0) | 0 (0.0) | 0 (0.0) | 0 (0.0) | 0 (0.0) | <0.001 |
| Unacceptable | 0 (0.0) | 0 (0.0) | 0 (0.0) | 0 (0.0) | 0 (0.0) |  |
| Mixed feelings | 15 (6.2) | 25 (29.4) | 34 (26.8) | 24 (27.9) | 98 (18.2) |  |
| Acceptable | 108 (45.0) | 45 (52.9) | 57 (44.9) | 29 (33.7) | 239 (44.4) |  |
| Strongly acceptable | 117 (48.8) | 15 (17.6) | 36 (28.3) | 33 (38.4) | 201 (37.4) |  |
| **Retrospective (n)** | **201** | **82** | **113** | **80** | **476** |  |
| Strongly unacceptable | 0 (0.0) | 0 (0.0) | 0 (0.0) | 0 (0.0) | 0 (0.0) | <0.001 |
| Unacceptable | 0 (0.0) | 0 (0.0) | 0 (0.0) | 0 (0.0) | 0 (0.0) |  |
| Mixed feelings | 17 (8.5) | 0 (0.0) | 39 (34.5) | 14 (17.5) | 70 (14.7) |  |
| Acceptable | 65 (32.3) | 18 (22.0) | 51 (45.1) | 28 (35.0) | 162 (34.0) |  |
| Strongly acceptable | 119 (59.2) | 64 (78.0) | 23 (20.4) | 38 (47.5) | 244 (51.3) |  |

### Factors associated with HCW’ acceptability

Compared to large or small PHC, working in a medium-sized PHC was positively associated with highest HCW’ prospective acceptability pre-PO training (adjusted OR (aOR)= 22.2, CI95%: 1.19-4.15, p=0.012), but no factor was found to be associated with the HCW’ prospective acceptability post-PO training (Table 3A-B). The e-IMCI support (aOR=2.82, CI95%: 1.58-5.03, p<0.001) and having been trained to use PO (aOR=2.48, CI95%: 1.64-3.74, p<0.001) were positively associated with highest HCW’ concurrent acceptability (Table 3C). Finally, being an “Assistant Nurse” (aOR=4.2, CI95%: 1.29-13.71, p=0.017) compared to a “Midwife” and having been trained to use PO were positively associated with HCW’ retrospective acceptability (Table 3D). Overall, neither age, sex nor years of experience were associated with highest HCW’ acceptability in the four surveys.

**Table 3.**
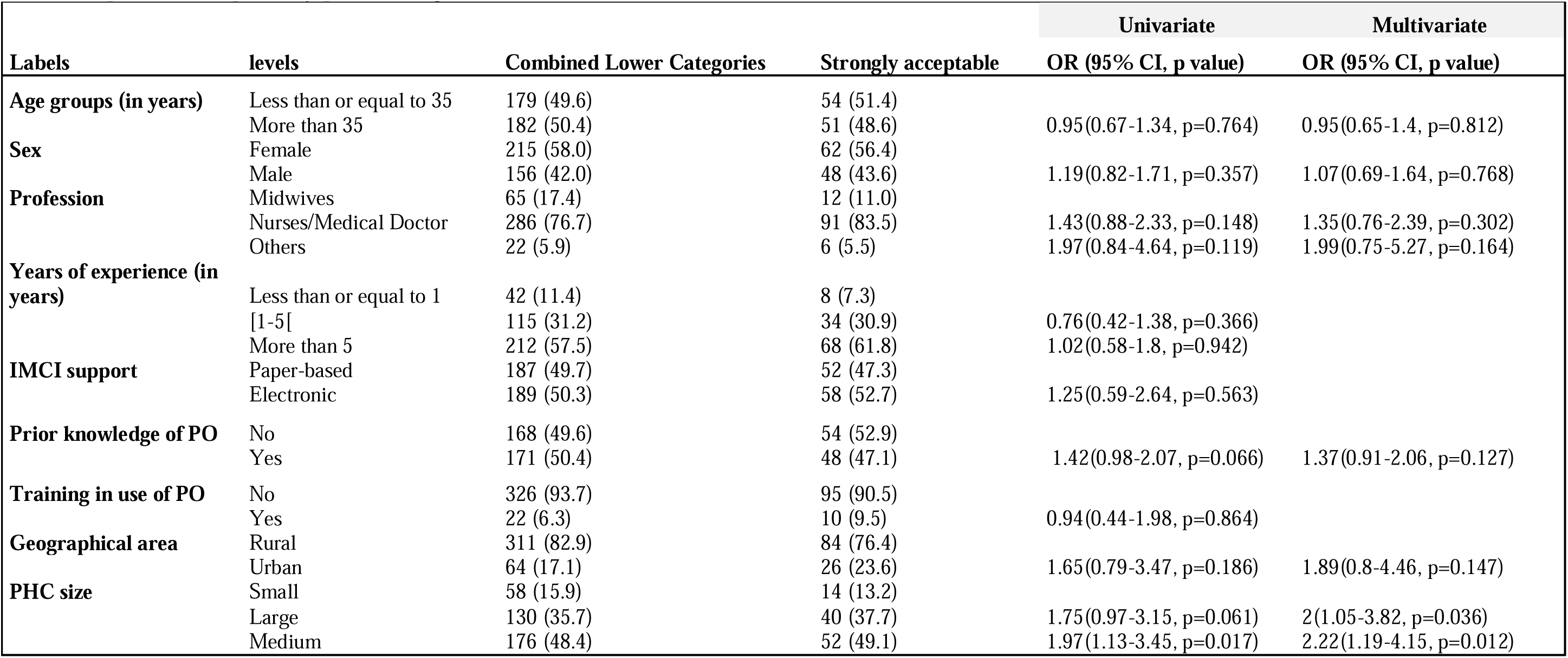
Factors associated to HCW’ strongly acceptability to PO use versus lower categories according to the time of data collection: Ordinal logistic regression modelling. 3A. Prospective acceptability pre-training.

**3B.** Prospective acceptability post-training.

3B - Prospective acceptability post-training
| Labels | levels | Combined Lower Categories | Strongly acceptable | Univariate | Multivariate |
| --- | --- | --- | --- | --- | --- |
|  |  |  |  | OR (95% CI, p value) | OR (95% CI, p value) |
| Age groups (in years) | Less than or equal to 35 | 143 (52.6) | 87 (47.3) |  |  |
|  | More than 35 | 129 (47.4) | 97 (52.7) | 1.05(0.74-1.5, p=0.782) | 1.01(0.71-1.45, p=0.938) |
| Sex | Female | 161 (57.1) | 112 (59.3) |  |  |
|  | Male | 121 (42.9) | 77 (40.7) | 1.24(0.84-1.82, p=0.277) | 1.29(0.86-1.92, p=0.215) |
| Profession | Midwives | 47 (16.7) | 29 (15.3) |  |  |
|  | Nurses/Medical Doctor | 220 (78.0) | 147 (77.8) | 1.17(0.71-1.92, p=0.542) |  |
|  | Others | 15 (5.3) | 13 (6.9) | 1.51(0.62-3.69, p=0.363) |  |
| Years of experience (in years) | Less than or equal to 1 | 28 (10.1) | 22 (11.6) |  |  |
|  | [1-5[ | 99 (35.6) | 46 (24.2) | 0.64(0.35-1.19, p=0.161) |  |
|  | More than 5 | 151 (54.3) | 122 (64.2) | 0.83(0.46-1.51, p=0.543) |  |
| IMCI support | Paper-based | 147 (45.0) | 116 (55.2) |  |  |
|  | Electronic | 180 (55.0) | 94 (44.8) | 0.59(0.27-1.25, p=0.168) | 0.71(0.29-1.77, p=0.466) |
| Geographical area | Rural | 239 (84.5) | 146 (76.0) |  |  |
|  | Urban | 44 (15.5) | 46 (24.0) | 1.85(0.84-4.09, p=0.127) | 1.84(0.7-4.82, p=0.215) |
| PHC size | Small | 75 (23.7) | 36 (17.6) |  |  |
|  | Large | 90 (28.5) | 79 (38.7) | 1.42(0.78-2.56, p=0.247) |  |
|  | Medium | 151 (47.8) | 89 (43.6) | 1.09(0.62-1.9, p=0.765) |  |

**3C.**
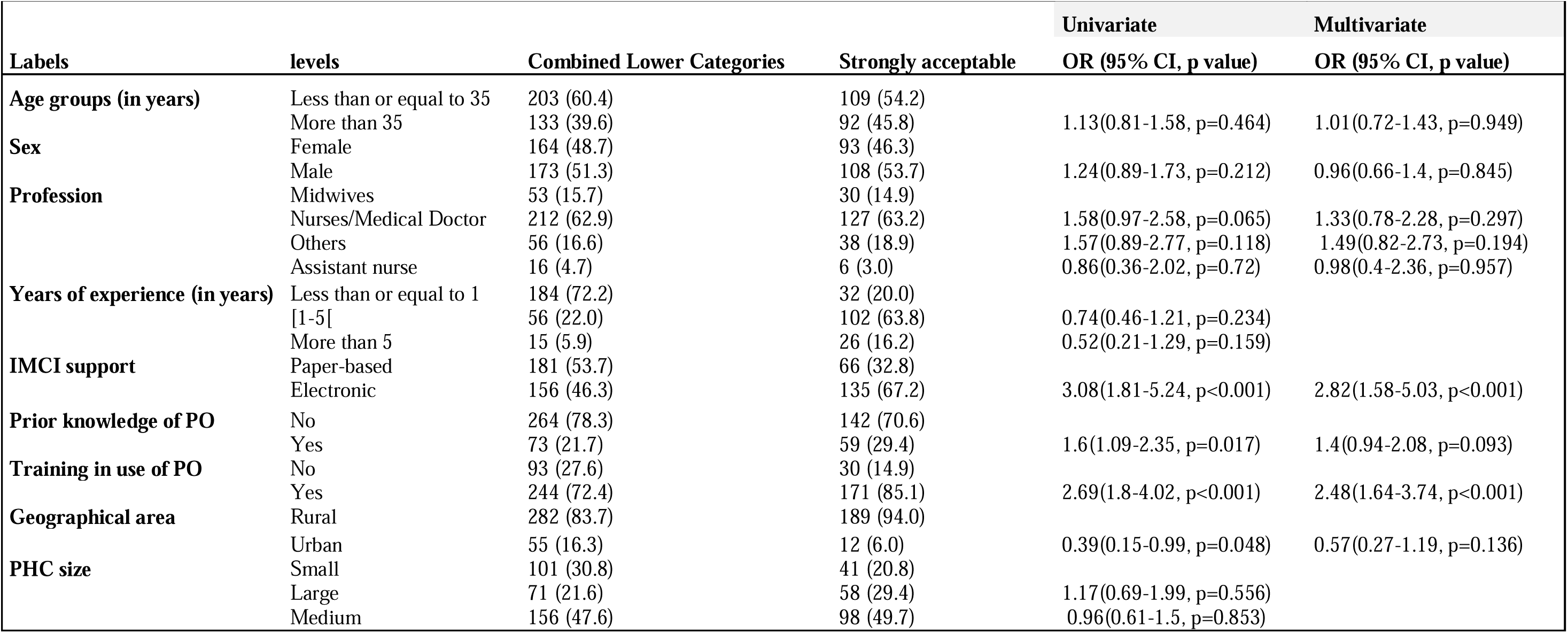
Concurrent acceptability.

**3D.** Retrospective acceptability.

3D - Retrospective acceptability
| Labels | levels | Combined Lower Categories | Strongly acceptable | Univariate | Multivariate |
| --- | --- | --- | --- | --- | --- |
|  |  |  |  | OR (95% CI, p value) | OR (95%CI, p value) |
| Age groups (in years) | Less than or equal to 35 | 138 (59.5) | 116 (47.5) |  |  |
|  | More than 35 | 94 (40.5) | 128 (52.5) | 1.37(0.94-1.99, p=0.103) | 1.24(0.83-1.83, p=0.292) |
| Sex | Female | 110 (47.4) | 108 (44.3) |  |  |
|  | Male | 122 (52.6) | 136 (55.7) | 1.43(0.97-2.11, p=0.067) | 1.16(0.76-1.77, p=0.505) |
| Profession | Midwives | 25 (10.8) | 10 (4.1) |  |  |
|  | Nurses/Medical Doctor | 142 (61.2) | 169 (69.3) | 2.56(1.26-5.18, p=0.009) | 2.03(0.95-4.31, p=0.067) |
|  | Others | 58 (25.0) | 50 (20.5) | 2.12(0.97-4.6, p=0.058) | 2.13(0.96-4.7, p=0.061) |
|  | Assistant nurse | 7 (3.0) | 15 (6.1) | 3.97(1.27-12.45, p=0.018) | 4.2(1.29-13.71, p=0.017) |
| Years of experience (in years) | Less than or equal to 1 | 154 (75.5) | 43 (18.5) |  |  |
|  | [1-5[ | 37 (18.1) | 140 (60.3) | 1.18(0.69-2.03, p=0.543) |  |
|  | More than 5 | 13 (6.4) | 49 (21.1) | 0.83(0.33-2.13, p=0.702) |  |
| IMCI support | Paper-based | 111 (47.8) | 115 (47.1) |  |  |
|  | Electronic | 121 (52.2) | 129 (52.9) | 1.05(0.33-3.29, p=0.938) |  |
| Prior knowledge of PO | No | 183 (78.9) | 186 (76.2) |  |  |
|  | Yes | 49 (21.1) | 58 (23.8) | 1.44(0.92-2.28, p=0.114) | 1.26(0.77-2.04, p=0.355) |
| Training in use of PO | No | 39 (16.8) | 39 (16.0) |  |  |
|  | Yes | 193 (83.2) | 205 (84.0) | 2.66(1.54-4.62 p<0.001) | 2.56(1.44-4.53, p=0.001) |
| Geographical area | Rural | 218 (94.0) | 199 (81.6) |  |  |
|  | Urban | 14 (6.0) | 45 (18.4) | 12.83(2.16-76.05, p=0.005) | 4.2(0.91-19.39, p=0.066) |
| PHC size | Small | 78 (33.9) | 50 (21.2) |  |  |
|  | Large | 47 (20.4) | 62 (26.3) | 1.44(0.77-2.69, p=0.259) | 1.23(0.64-2.36, p=0.533) |
|  | Medium | 105 (45.7) | 124 (52.5) | 1.64(1-2.7, p=0.051) | 1.57(0.94-2.61, p=0.084) |

### Acceptability by dimension

Table 4 summarizes the quantitative acceptability of HCW by dimension, and key qualitative elements are presented in Appendix 4.

**Table 4.**
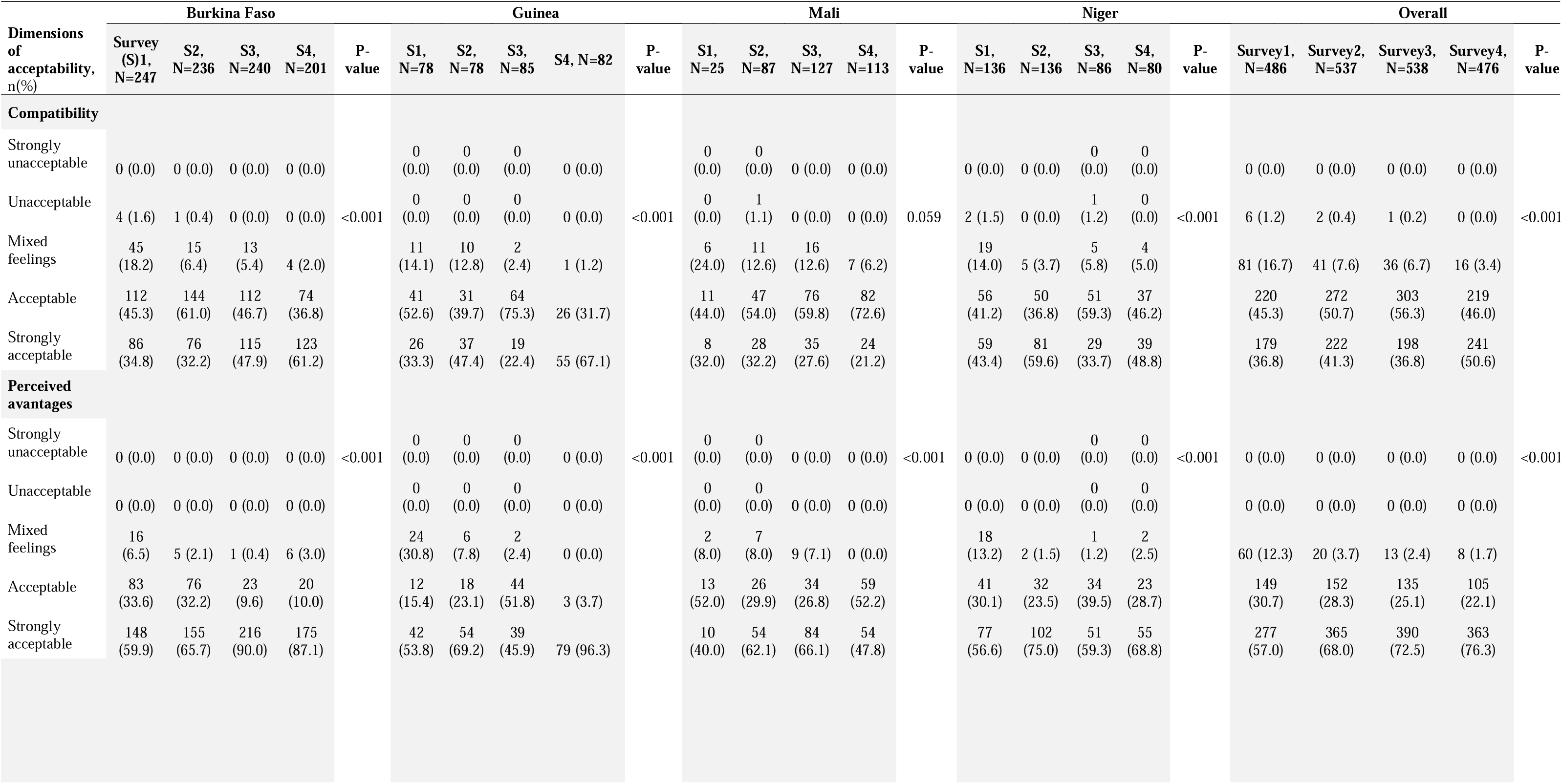
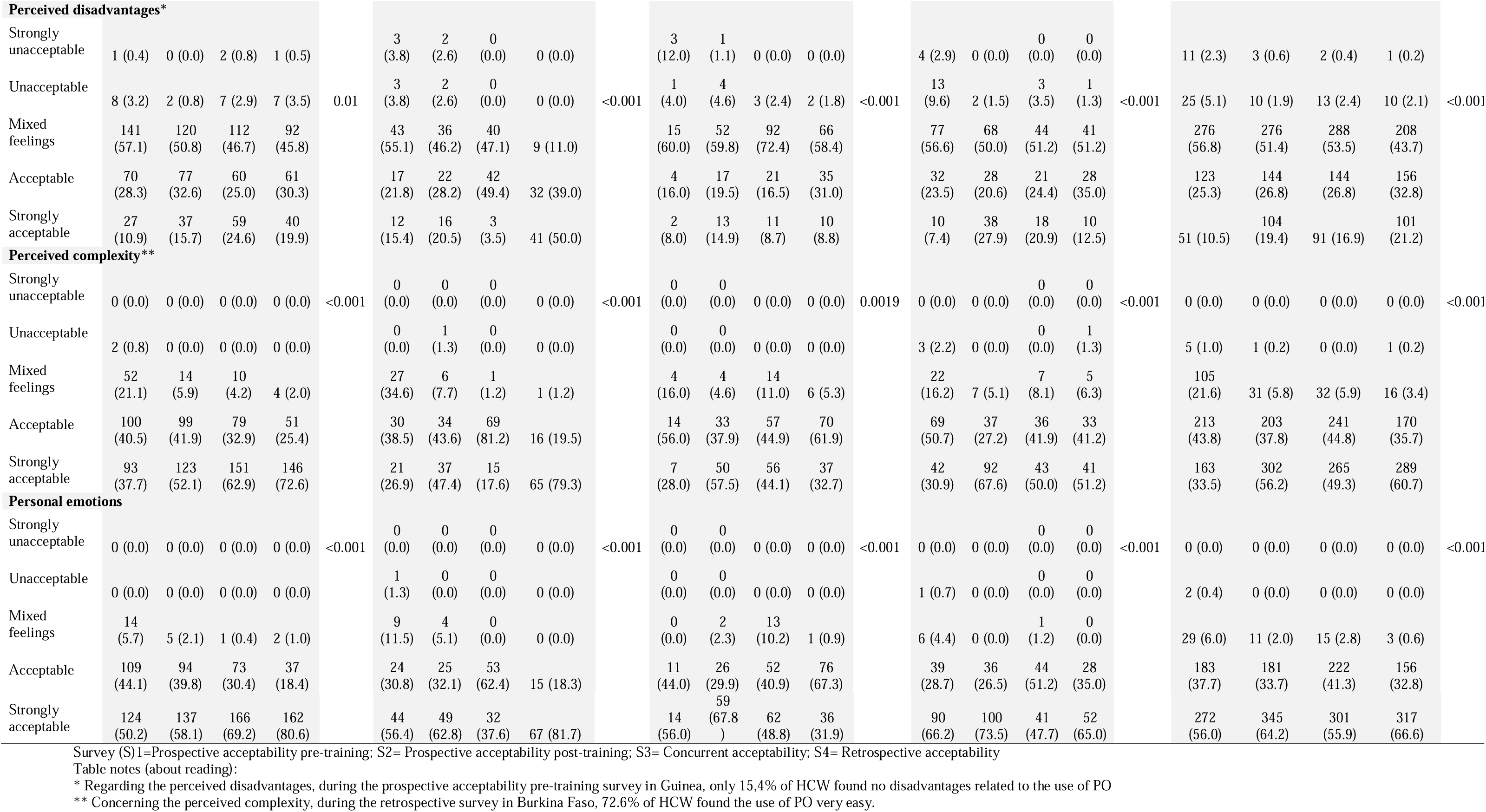
Evolution of Health Care Workers’ acceptability over time surveys by dimensions except social influence dimension (presented in appendix 5) according to the Likert scale, overall and by country (N=202 AIRE Primary Health Centres).

### Compatibility

The overall proportion of HCW who found the PO strongly compatible with their context increase from 36.8% to 50.6% between the pre-training and retrospective survey (p-value<0.001). This trend was consistent across all countries except Mali, where the proportion of HCW decreased from 32.0% to 21.2%. However, this was offset by HCW finding the PO compatible (but not strongly).

Indeed, HCW felt that the PO meets the crucial need to improve their PHC’s technical capabilities, providing reliable equipment at a decentralized level for better patient care. This feeling was expressed during both qualitative acceptability phases: the concurrent (T1) and retrospective (T2) surveys.

The PO is seen as compatible and complementary to IMCI and existing consultation practices. However, a few HCW in Mali noted that consultation documents, such as paper registers, were not fully adapted for PO use, as there is no specific space for recording saturation levels. Contextual factors, such as the existing work organisation within the PHC, also influence PO’s compatibility. In Burkina Faso, it was mentioned that using a single consultation room for both children and adults could complicate PO integration. In Mali and Niger, some PHC reorganized their working areas by separating children’s and adult consultation rooms to integrate the PO more effectively.

HCW also noted the PO’s good compatibility reflected in its acceptance by communities, as it does not contradict any values or beliefs. Some parents even requested its use:

> *”The mother will tell you that you didn’t check the child with the machine (…) According to her, you didn’t consult her child properly. You’re going to explain it and then take it just to reassure her”.* (Burkina Faso, T2).

### Perceived complexity

We noted an increase in the proportion of HCW who perceived the PO as very easy to use between the first and last data collection (p-value<0.001). Nevertheless, this improvement is not linear, and during the concurrent data collection, the number of HCW finding the PO very easy to use significantly decreased in all countries except Burkina Faso.

According to the interviews, the perceived ease of use of the PO for HCW depends on whether they have been trained. Those who received training found it easy to use. However, staff who did not receive formal training found the PO much more complicated, especially in Guinea. This is consistent with the quantitative results, where only 17.6% of Guinean HCW found the PO very easy to use during the concurrent phase. Feedback sessions from trained HCW were supposed to be organized but were often insufficient or not held. In some cases, untrained HCW could use the PO and record oxygen saturation but didn’t know what the tool was for or how to interpret the data, leaving interpretation to the PHC’ head.

> *”I don’t know the parameters, if it’s high or not…if the PO shows 99%, I don’t know if it’s normal, because sometimes you find it’s 94%, I don’t know the difference, if it’s normal or if it’s not normal…a number appears and that’s what I write”.* (Guinea, T1)

This situation improved over time, as supervision played a significant role in helping untrained HCW master the PO. Finally, incorporating the PO into the IMCI algorithm sometimes complicated its use, as the algorithm was not always well-used before the project.

However, issues related to this inexperience were much less frequent in the retrospective phase.

### Perceived advantages

Acceptability scores for this dimension are very high, particularly in the retrospective study in Burkina Faso (87.1%); and in Guinea (96.3%) despite a drop to 45.9% during concurrent data collection. These rates were lower in Mali (47.8%) and Niger (68.8%), but the proportion of those with mixed feelings was extremely low in all countries. According to the qualitative data, HCW generally found many advantages in using the PO during the two rounds of data collection. First, decentralizing diagnostic improves care from the beginning of the process and HCW are pleased to access tools previously reserved for higher levels of the health system.

The PO is also perceived as a tool that guides their practices and helps them make decisions. Better diagnosis avoids unnecessary trips between home and PHC and improves the hospital referral process when necessary.

> *”Before the PO, doctors were slow to make decisions to refer cases. They kept them at the center, believing that they could find a solution. But with the PO, if it’s a case that needs to be referred, we know right away and don’t take long to decide.”* (Mali, T1)

The PO allows HCW to be more confident about their diagnoses. Good saturation reassures them, while low saturation facilitates rapid decision-making. Several HCW also highlighted that new equipment like the PO improves their credibility and the PHC’s image in the community.

### Perceived disadvantages

We noted an increase in the proportion of HCW who perceived disadvantages in using the PO; varying overall from 10.5% to 21.2% (p-value<0.001), and markedly observed in Guinea. The main difficulty identified from interviews was using the PO with agitated children, as it can be time-consuming or impossible. This agitation is often linked to their fear of being pricked, as with the malaria rapid diagnostic test (mRDT), since the sensor is placed on the finger. HCW employ strategies to calm the child, such as asking the mother for help, demonstrating on their own finger, or taking the saturation before using the mRDT.

Another perceived disadvantage was the increased workload due to integrating PO use within the IMCI algorithm. HCW must take a lot of parameters (weight, temperature, arm circumference, etc). However, this impression was partly due to HCW’s inexperience with the PO and IMCI. With time and practice, they perceived the PO much less as an additional workload.

An increased number of tests to diagnose children can lead to longer consultation times, which is challenging when human resources are limited or during busy periods in the PHC; and sometimes lead to patient dissatisfaction.

> *”Children are often agitated, which makes it difficult for the PO to give the result, and that makes you late. The people waiting yell at you, saying you’re slow.”* (Mali, T2)

However, these difficulties were more emphasized during the first qualitative data collection. Improved control of the PO, recognition of its importance, and community awareness-raising reduced complaints about the increase in consultation time.

Some HCW view the longer consultation time positively, as it allows for better identification of health problems, a perception influenced by contextual factors such as the busyness of the PHC.

Another challenge is the high turnover of HCW. Proper use of the device requires training, and frequent staff changes mean constant retraining is necessary. Finally, the low socio-economic resources of the population can lead to difficulties for HCW because CF sometimes refuse referrals. This is often due to the fear of hospital referrals, which highlight the severity of their child’s condition and incur expenses they often cannot afford.

### Personal emotions

Positive or very positive feelings about the PO remained high in all four countries during the four surveys. The proportions of HCW with very positive feelings increased from 50.2% in the first survey to 80.6% in the last survey in Burkina Faso and from 56.4% to 81.7% in Guinea. However, these proportions decreased in the other two countries (p-value<0.001). Nevertheless, the declines were offset by those finding the PO “acceptable,” with only three people having mixed feelings in the last data collection compared to 29 in the first survey.

This was reflected in the interviews; overall, HCW expressed positive feelings about the PO over time. It improved their self-confidence, as they felt more confident of their diagnoses.

> *”I feel at ease, it gives me confidence in what I’m doing, because I know that the device isn’t going to lie to me. It gives me the correct result.”* (Mali, T2)

Receiving new technology to assist them in their daily work also brings satisfaction and joy. They feel rewarded by having access to a device that is rare in these contexts. However, a few HCW remain apprehensive about their ability to use the tool correctly or whether the PO provides reliable data. This concern is linked to specific conditions for using the PO (clean fingers, no cold hands, etc.) and whether they received adequate training.

### Social influence

Most HCW interviewed felt that others couldn’t influence their opinion of the PO because they understood its usefulness and importance. This sentiment was reaffirmed during the retrospective phase.

However, some HCW, particularly in Guinea, stated that a contrary opinion from their superior in the PHC or health district team could influence their use of the tool, even if they wished to use it.

Some PHC’ manager recognize their influence and understand they must model the desired behavior.

> *”As the manager, if you consult without using it, you should know that it will never be used. So, every time you lead by example, you have to do it so that others see you’re serious about using it.”* (Burkina Faso, T1)

The long-term involvement of project teams through supervision also contributed to the positive perception of the PO, reinforcing the importance of using it to HCW.

In the quantitative data, almost no HCWs reported negative influences from health authorities, colleagues, or PHC heads during the concurrent acceptability survey. Positive reactions from children, mothers, or caregivers were common, though a minority reported negative influences in Burkina Faso and Mali. Negative influences decreased in these two countries in the retrospective survey but increased by 10.0% in Guinea and 15.4% in Niger (p-value=2.2e-05) (Appendix 5).

### CF’ qualitative acceptability by dimension

Appendix 6 highlights key elements of CF’ qualitative acceptability by dimension.

### Compatibility

Generally, CF found the PO compatible with existing values, customs, and practices. The PO’s noninvasive nature facilitates its acceptance by CF. Moreover, the PO complements existing practices without altering them. HCW also reported that patients visiting PHC are rarely hesitant about new equipment.

### Perceived complexity

Few CFs had any information or knowledge about the PO’s usefulness and the project’s objectives. Several HCW emphasized the importance of raising awareness among CF to help them understand and accept the PO. However, during consultation observations, we noted that very little or no information was provided to CF about the device’s purpose and usage. CF often avoid questioning HCW due to several barriers, like language and health literacy.

> *”All I see is a mobile phone, which I think is for consultation purposes, but I don’t know what it’s for. When it was placed on my child, it displayed something, but I can’t read to understand. And the nurse doesn’t speak Zarma so we can’t understand each other.”* (Niger)

### Perceived advantages

CF believe that having new tools at the peripheral level improve the quality of care and treatment, enabling the detection of illnesses that couldn’t be identified before.

> *”Human beings have their limits, so to be more efficient at work, they need a device that complements these human limits. The machine quickly produces the result; it shows you straight away what your patient’s problem is.”* (Mali)

The PO optimizes care pathways. It helps avoid round trips to the PHC and quickly identifies whether a child needs to be referred to the hospital.

However, the real benefits of the tool can sometimes be overestimated, with some CF perceiving the PO as capable of detecting or even curing all diseases. Other elements, such as donations of medicines and follow-up care for children (as part of another research component), may also have contributed to community acceptability.

### Perceived disadvantages

Many CF, like HCW, felt that consultation times had increased. While some found this disruptive, especially on market days, others felt it was the most important thing if it was necessary for the child’s well-being.

> *”When they conduct the consultation with the new device (…) it’s more work and it delays the consultation a bit. But I understand that they’re doing something else that they weren’t doing before.”* (Guinea)

A few CF even noticed that waiting times had decreased since the introduction of the PO. In addition, as already mentioned by HCW, some CF noted their child’s reluctance towards the device and highlighted issues related to hospital referrals, mainly due to their costs.

### Personal emotions

Overall, CF expressed positive feelings about the PO, showing confidence in the device and the HCW’s ability to use it correctly. Many CF felt joy, either because their child’s health improved, or because they could understand the seriousness of their child’s condition thanks to the device.

> *”I felt nothing but joy in my heart. A joy in knowing that my daughter is being treated.”* (Niger)

A few parents expressed fears after seeing HCW struggle to use the device on their child or due to concerns about misreading affecting their child’s health. However, such issues were rare (few cases of refusal) and raising awareness of the PO’s usefulness usually alleviated these fears.

### Social influence

CF stated that they relied heavily on the advice of HCW, who are seen as trusted health experts. In any case, they often have no other option but to take their child to the PHC in case of a health problem and accept what will be offered there.

## Discussion

To our knowledge, this study is the first comprehensive assessment of the acceptability of routine PO use integrated into IMCI guidelines at the PHC level in West Africa. Conducted in diverse settings, our research used a robust conceptual framework and mixed methods over time, significantly enhancing our understanding of PO acceptability.

The overall acceptability is high, despite variations over time and between countries. The integration of quantitative and qualitative data reveals convergences. The results related to the different dimensions overlap between the qualitative and the quantitative data. Moreover, the factors associated with higher acceptability at the quantitative level, such as the influence of PHC size, use of e-IMCI, profession, and the crucial role of training in PO use, also emerged at the qualitative level. Interviews revealed that using the PO can be more complicated in busy centers due to extended consultation time. In PHC using e-IMCI, HCW were more familiar with IMCI and electronic devices, reducing the perceived workload of PO use. Training and supervision were crucial factors in the evolution of acceptability over time. Training often prioritizes individuals for political and institutional reasons [30], typically involving heads of PHC and their deputies, who may not always engage directly in the tasks they were trained for. In Guinea, where PO was little known before the project (only 9.4% of HCW), intense supervision significantly enhanced acceptability over time, allowing HCW to ask questions, receive feedback, and master the device.

Interviews also emphasized the importance, for acceptability, of the improvement of confidence in the diagnosis. This aligns with other studies, such as in India, where a PO increased HCW’s confidence in their decision-making regarding child management [19]. Another study found that introducing PO in Malawi and Bangladesh increased confidence, especially for frontline workers who often lack diagnostic tools and have less confidence in their clinical judgment [18]. This boost in confidence also led to increased hospital referrals in India [19]. Similarly, in the AIRE project, we observed a higher proportion of hypoxemic children being referred compared to severe cases without hypoxemia [31], as also observed in Malawi [32]. This is crucial given the challenges of diagnostic accuracy and correct referral from PHC [33]. The importance of good training, regular feedback and continuous support in using PO is also highlighted in several studies, both at the hospital level [8,16] and in decentralised settings [20].

The integration of quantitative and qualitative results did not reveal any divergent factors. However, some acceptability challenges were not specifically sought quantitatively but emerged qualitatively. Increased consultation times and workload, especially with agitated children, and difficulties with hospital referrals for children with hypoxemia were recurrent themes in interviews. The challenges related to child agitation are also reported in other studies [18]. Referral difficulties, even when HCW detects illness severity, are well known in these contexts [34,35]. A study in Burkina Faso showed that only 41.5% of referred patients visited the referral hospital within seven days after the HCW’s decision [36]. Oxygen is often unavailable at decentralized levels, which may discourage HCW if only the diagnosis of hypoxemia improves without access to treatment.

Finally, CF generally accepted PO with minimal resistance despite limited knowledge of its actual benefits. This aligns with other studies showing high acceptability among patients’ families and an increased trust in HCW and their diagnoses [18,19,37].

Our study also highlighted the importance of distinguishing between acceptability and use or participation, concepts often conflated in analyses [38,39]. Despite initial variability, PO uptake rates exceeded 90% in the last ten months, with no significant differences between countries. However, this uptake appears somewhat disconnected from acceptability rates. The project’s context, its benefits (equipment donations, medicines, incentives, etc.) and regular supervision funded by the project may compel HCW to report usage due to pressure for positive outcomes [40]. A comprehensive understanding of the device’s acceptability provides insights into its sustainability [14]. However, its long-term sustainability needs to be studied, given that the medical equipment supplied in this context does not generally last [41,42].

Our study has some limitations. Qualitative data were collected only in the research PHC, where HCW were more closely monitored, potentially skewing the representativeness and generalizability of results. Interview responses may reflect social desirability bias despite mitigation efforts [43], especially given the NGO-led project’s benefits. Separating opinions on PO from the broader project context, which inevitably impacts acceptability, is challenging. It is also necessary to interpret CF acceptability results considering the power dynamics and psychosocial pressures that may limit the expression of negative critiques from patients and their families [44]. In the quantitative study, high turnover made maintaining cohort follow-up difficult. Instead, we used repeated cross-sectional surveys, limiting our ability to track acceptability changes over time. Another limit was the uniform weighting of questions and dimensions in calculating acceptability scores, which may not reflect their actual impact. This article is the first attempt to test our conceptual framework empirically [23], and further research is needed to refine our methodology and potentially introduce weighting coefficients.

### Conclusion

We found that both HCW and CF generally accept the PO. While CF did not always understand the device’s utility, their overall perception was positive. HCW who received proper training had significantly higher acceptability. Perceived advantages of PO included improved childcare, more accurate diagnoses, and enhanced self-confidence. However, challenges such as repeated training needs, increased workload, longer consultation time, and referral difficulties to hospitals could impact the sustainability of PO use in West Africa.

## ACKNOWLEDGMENTS

We thank all the participants of this study, as well as the healthcare staff involved in the AIRE project. We also express our gratitude to the field project staff and the AIRE Research Study Group. Finally, we thank the Ministries of Health of the participating countries for their support.

Acknowledgements: Children, families, UNITAID and The AIRE Research Study Group:

**Country investigators**: Ouagadougou, Burkina Faso: S. Yugbaré Ouédraogo (PI), V. M. Sanon Zombré (CoPI), Conakry, Guinea: M. Sama Cherif (CoPI), I. S. Diallo (CoPI), D. F. Kaba, (PI). Bamako, Mali: A. A. Diakité (PI), A. Sidibé, (CoPI). Niamey, Niger: H. Abarry Souleymane (CoPI), F. Tidjani Issagana Dikouma (PI). **Research coordinators & data centers: Inserm U1295, Toulouse 3 University, France:** H. Agbeci (Int Health Economist), L. Catala (Research associate), D. L. Dahourou (Research associate), S. Desmonde (Research associate), E. Gres (PhD Student), G. B. Hedible (Int research project manager), V. Leroy (research coordinator), L. Peters Bokol (Int clinical research monitor), J. Tavarez (Research project assistant), Z. Zair (Statistician, Data scientist). **CEPED, IRD, Paris, France:** S. Louart (process manager), V. Ridde (process coordination). **Inserm U1137, Paris, France:** A. Cousien (Research associate). **Inserm U1219,** EMR271 IRD, **Bordeaux University, France**: R. Becquet (Research associate), V. Briand (Research associate), V. Journot (Research associate). **PACCI, CHU Treichville, Abidjan, Côte d’Ivoire**: S. Lenaud (Int data manager), C. N’Chot (Research associate), B. Seri (Supervisor IT), C. Yao (data manager supervisor). **Consortium NGOs partners: Alima-HQ (consortium lead), Dakar, Sénégal**: G. Anago (Int Monitoring Evaluation Accountability And Learning Officer), D. Badiane (Supply chain manager), M. Kinda (Director), D. Neboua (Medical officer), P. S. Dia (Supply chain manager), S. Shepherd (referent NGO), N. di Mauro (Operations support officer), G. Noël (Knowledge broker), K. Nyoka (Communication and advocacy officer), W. Taokreo (Finance manager), O. B. Coulidiati Lompo (Finance manager), M. Vignon (Project Manager). **Alima, Conakry, Guinea:** P. Aba (clinical supervisor), N. Diallo (clinical supervisor), M. Ngaradoum (Medical Team Leader), S. Léno (data collector), A. T. Sow (data collector), A. Baldé (data collector), A. Soumah (data collector), B. Baldé (data collector), F. Bah (data collector), K. C. Millimouno (data collector), M. Haba (data collector), M. Bah (data collector), M. Soumah (data collector), M. Guilavogui (data collector), M. N. Sylla (data collector), S. Diallo (data collector), S. F. Dounfangadouno (data collector), T. I. Bah (data collector), S. Sani (data collector), C. Gnongoue (Monitoring Evaluation Accountability And Learning Officer), S. Gaye (Monitoring Evaluation Accountability And Learning Officer), J. P. Y. Guilavogui (Clinical Research Assistant), A. O. Touré (Country health economist), J. S. Kolié (Country clinical research monitor), A. S. Savadogo (country project manager). **Alima, Bamako, Mali:** F. Sangala (Medical Team Leader), M. Traore (Clinical supervisor), T. Konare (Clinical supervisor), A. Coulibaly (Country health economist), A. Keita (data collector), D. Diarra (data collector), H. Traoré (data collector), I. Sangaré (data collector), I. Koné (data collector), M. Traoré (data collector), S. Diarra (data collector), V. Opoue (Monitoring Evaluation Accountability And Learning Officer), F. K. Keita (medical coordinator), M. Dougabka (Clinical research assistant then Monitoring Evaluation Accountability And Learning Officer), B. Dembélé (data collector then Clinical research assistant), M. S. Doumbia (country health economist), G. D. Kargougou (country clinical research monitor), S. Keita (country project manager). **Solthis-HQ, Paris**: S. Bouille (NGO referent), S. Calmettes (NGO referent), F. Lamontagne (NGO referent). **Solthis, Niamey:** K. H. Harouna (clinical supervisor), B. Moutari (clinical supervisor), I. Issaka (clinical supervisor), S. O. Assoumane (clinical supervisor), S. Dioiri (Medical Team Leader), M. Sidi (data collector), K. Sani Alio (Country supply chain officer), S. Amina (data collector), R. Agbokou (Clinical research assistant), M. G. Hamidou (Clinical Research Assistant), S. M. Sani (Country health economist), A. Mahamane, Aboubacar Abdou (data collector), B. Ousmane (data collector), I Kabirou (data collector), I. Mahaman (data collector), I Mamoudou (data collector), M. Baguido (data collector), R. Abdoul (data collector), A. Sahabi (data collector), F. Seini (data collector), Z. Hamani (data collector), L-Y B Meda (Country clinical research monitor), Mactar Niome (country project manager), X. Toviho (Monitoring Evaluation Accountability And Learning Officer), I. Sanouna (Monitoring Evaluation Accountability And Learning Officer), P. Kouam (program officer). **Terre des hommes-HQ, Lausanne:** S. Busière (NGO referent), F. Triclin (NGO referent). **Terre des hommes, BF:** A. Hema (country project manager), M. Bayala (IeDA IT), L. Tapsoba (Monitoring Evaluation Accountability And Learning Officer), J. B. Yaro (Clinical reearch assistant), S. Sougue (Clinical reearch assistant), R. Bakyono (Country health economist), A. G. Sawadogo (Country clinical research monitor), A. Soumah (data collector), Y. A. Lompo (data collector), B. Malgoubri (data collector), F. Douamba (data collector), G. Sore (data collector), L. Wangraoua (data collector), S. Yamponi (data collector), S. I. Bayala (data collector), S. Tiegna (data collector), S. Kam (data collector), S. Yoda (data collector), M. Karantao (data collector), D. F. Barry (Clinical supervisor), O. Sanou (clinical supervisor), N. Nacoulma (Medical Team Leader), N. Semde (clinical supervisor), I. Ouattara (Clinical supervisor), F. Wango (clinical supervisor), Z. Gneissien (clinical supervisor), H. Congo (clinical supervisor). **Terre des hommes, Mali:** Y. Diarra (clinical supervisor), B. Ouattara (clinical supervisor), A. Maiga (data collector), F. Diabate (data collector), O. Goita (data collector), S. Gana (data collector), S. Diallo (data collector), S. Sylla (data collector), D. Coulibaly (Tdh project manager), N. Sakho (NGO referent). **Country SHS team: Burkina Faso**: K. Kadio (consultant and research associate), J. Yougbaré (data collector), D. Zongo (data collector), S. Tougouma (data collector), A. Dicko (data collector), Z. Nanema (data collector), I. Balima (data collector), A. Ouedraogo (data collector), A. Ouattara (data collector), S. E. Coulibaly (data collector). **Guinea**: H. Baldé (consultant and research associate), L. Barry (data collector), E. Duparc Haba (data collector). **Mali**: A. Coulibaly (consultant and research associate), T. Sidibe (data collector), Y. Sangare (data collector), B. Traore (data collector), Y. Diarra (data collector). **Niger**: A. E. Dagobi (consultant and research associate), S. Salifou (data collector), B. Gana Moustapha Chétima (data collector), I. H. Abdou (data collector)

## FOOTNOTES

### Contributors

SL, HGB, VL and VR conceptualised the research. HB, AC, AED, KK and DN conducted data collection with the help of the AIRE research study group. SL, HGB, HB, AC, AED, KK, and ZZ realised the data analysis. SL and HGB wrote the first draft of this article. All authors were involved in data interpretation and review of the final manuscript.

## Ethics approval and consent to participate

The AIRE research protocol, the information notice (translated in vernacular languages), the written consent form and any other relevant document have been submitted to each national ethics committee, to the Inserm Institutional Evaluation Ethics Committee (IEEC) and to the WHO Ethics Review Committee (WHOLJERC). All the aforementioned ethical committees reviewed and approved the protocol and other key documents (Comité d’Ethique pour la Recherche en Santé (CERS), Burkina Faso n°2020–4LJ070; Comité National d’Ethique pour la Recherche en Santé (CNERS), Guinea n°169/CNERS/21; Comité National d’Éthique pour la Santé et les Sciences de la vie (CNESS), Mali n°127/MSDSLJCNESS; Comité National d’Ethique pour la Recherche en Santé (CNERS) Niger n°67/2020/CNERS; Inserm IEEC n°20–720; WHOLJERC n° ERC.0003364). This study has been retrospectively registered by the Pan African Clinical Trials Registry on June 15^th^, 2022, under the following Trial registration number: PACTR202206525204526.

## Data Availability Statement

The datasets generated and analysed during the current study are not publicly available. Access to processed deidentified participant data will be made available to any third Party after the publication of the main AIRE results stated in the Pan African Clinical Trial Registry Study statement (PACTR202206525204526, registered on 06/15/2022), upon a motivated request (concept sheet), and after the written consent of the AIRE research coordinator (Valériane Leroy, Inserm U1295 Toulouse, France, orcid.org/0000-0003-3542-8616) obtained after the approval of the AIRE publication committee, if still active.

---

**Appendix 1.** Example of concurrent acceptability – questions by dimension. All questions were based on a Likert scale ranging from strongly disagree to strongly agree.

| Dimensions | Questions |
| --- | --- |
| Compatibility | 1. Your PHC has all the necessary equipment to use and integrate the PO correctly into care routines.<br>2. PO is a tool that can meet the needs you encounter in your clinical practice.<br>3. PO can meet the needs of the PHC in which you work.<br>4. PO is a tool that fits well into the context (organizational, social, economic, etc.) of your PHC. |
| Perceived complexity | 1. PO is difficult to use.<br>2. You understand how using PO during consultations can improve the care of children under five.<br>3. You are confident in your ability to use the PO correctly during consultations.<br>4. The procedures for using PO are clear. |
| Perceived benefits | 1. PO will improve the system for referring serious cases to hospitals.<br>2. PO can improve diagnosis of children under five.<br>3. PO can reduce mortality from respiratory difficulties.<br>4. The use of PO is not useful. |
| Perceived disadvantages | 1. It is difficult to integrate the use of PO into IMCI.<br>2. The use of PO during IMCI consultations complicates your work.<br>3. Using the PO interferes with the other tasks you have to perform during the consultation.<br>4. PO increases consultation time.<br>5. PO causes work overload. |
| Personal emotions | 1. You have a positive feeling towards the PO.<br>2. You are willing to use PO during your consultations. |
| Social influence | 1. Your acceptability of the PO may be influenced by the recommendations of your PHC manager.*<br>2. Your acceptability of the PO may be influenced by the reaction of children and/or their families to the tool.*<br>3. Your acceptability of the PO may be influenced by the opinion of the Ministry of Health/health authorities regarding this tool.*<br>4. Your acceptability of the PO may be influenced by your colleagues' opinion of the tool.*<br>* For each of these questions, if the respondents answered "agree" or "strongly agree", they were then asked:<br>If yes, did they influence:<br>☐ Positively your opinion of the PO<br>☐ Negatively your opinion of the PO<br>☐ Have not influenced your opinion so far |

**Appendix 2.** Composite reliability test per acceptability’ dimension.

| **Acceptability' dimensions** | **Composite Reliability (CR)** |  |  |  |
| --- | --- | --- | --- | --- |
|  | **Survey 1** | **Survey 2** | **Survey 3** | **Survey 4** |
| Compatibility | 0.70 | 0.66 | 0.70 | 0.73 |
| Perceived advantages | 0.61 | 0.60 | 0.77 | 0.74 |
| Perceived disadvantages | 0.79 | 0.71 | 0.75 | 0.77 |
| Perceived complexity | 0.66 | 0.60 | 0.72 | 0.72 |
| Personal emotions | 0.70 | 0.63 | 0.81 | 0.78 |

Survey 1: Prospective acceptability pre-training
Survey 2: Prospective acceptability post-training
Survey 3: Concurrent acceptability
Survey 4: Retrospective acceptability

**Apendix 3.**
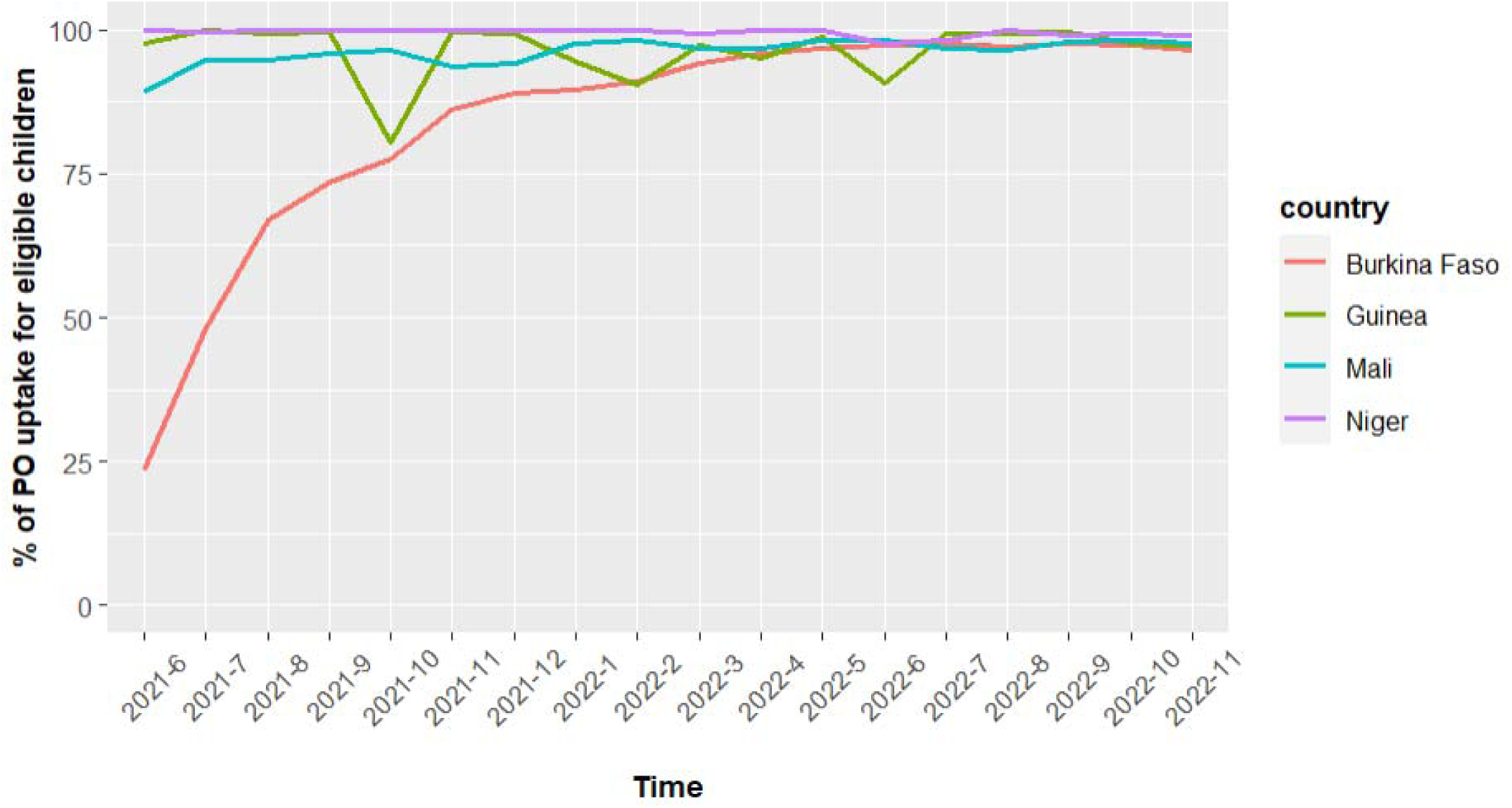
Evolution of the monthly Pulse Oximeters uptake (%) for eligible children (all IMCI children except children aged from 2-59 months, classified as green cases without respiratory signs) in the 202 AIRE Primary Health Centres by country, June 2021-December 2022.

**Appendix 4.** Key elements related to qualitative HCW’ acceptability.

| Dimensions | Key elements |
| --- | --- |
| Compatibility | • Meets the need to improve the PHC's technical platform<br>• Complementary with IMCI<br>• Not compatible with certain supports, such as IMCI registers (Mali)<br>• Compatibility may depend on the pre-existing organization of work within the PHC and the number of consultation rooms |
| Perceived complexity | • Quite easy to use with training<br>• Untrained HCW may have difficulty using the device and interpreting the results<br>• Familiarity with technologies such as smartphones can reduce perceived complexity<br>• Integration with IMCI can sometimes be difficult for HCW<br>• Supervision and practical experience with the device help reduce perceived complexity over time |
| Perceived advantages | • Contributes to the availability of diagnostic tools at a decentralized level<br>• Helps with quick decision-making and reduces unnecessary trips to the PHC<br>• Increases HCW confidence in their diagnosis<br>• Enhances the image of the PHC and HCW among the population<br>• Allows HCW to acquire new skills |
| Perceived disadvantages | • Difficult to use with agitated children (fear of being pricked, like with the mRDT)<br>• Initial increase in workload, particularly related to the integration of the device into IMCI (perception decreases over time)<br>• Increased consultation time causes difficulties and complaints, especially with high patient volumes or low HCW availability<br>• Difficulties for patients in accepting and paying for referrals<br>• High turnover but proper use of the device requires training |
| Personal emotions | • Improves HCW self-confidence and confidence in their diagnosis (reassuring)<br>• Joy related to the device's ability to help HCW and provide benefits<br>• Sometimes a bit of fear due to the risk of misinterpreting the data (Mali) |
| Social influence | • HCW' opinions are generally not influenced as they recognize the device's importance<br>• Patients' opinions have little influence<br>• Supervisors at the PHC or district level can sometimes influence their opinions |

**Appendix 5.** Description of social influence acceptability’ dimension of Health Care Workers during the concurrent and retrospective acceptability surveys, (N=202 AIRE Primary Health Centres).

|  | Concurrent acceptability |  |  |  | P-value | Retrospective acceptability |  |  |  | P-value |
| --- | --- | --- | --- | --- | --- | --- | --- | --- | --- | --- |
|  | Burkina<br>Faso<br>N=240 | Guinea<br>N=85 | Mali<br>N=127 | Niger<br>N=86 |  | Burkina<br>Faso<br>N=201 | Guinea<br>N=82 | Mali<br>N=113 | Niger<br>N=80 |  |
| **Opinion of the ministry of health/health authorities** | N=131 | N=29 | N=58 | N=44 |  | N=125 | N=77 | N=56 | N=41 |  |
| Has not influenced your opinion about PO yet, n (%) | 33 (25.2) | 24 (82.8) | 22 (37.9) | 4 (9.1) |  | 1 (0.8) | 11 (14.3) | 30 (53.6) | 3 (7.3) |  |
| Negative influence on your opinion about PO, n (%) | 1 (0.8) | 0 (0.0) | 0 (0.0) | 0 (0.0) | 1.76e-10 | 1 (0.8) | 0 (0.0) | 0 (0.0) | 1 (2.4) | 4.34e-17 |
| Positive influence on your opinion about PO, n (%) | 97 (74.0) | 5 (17.2) | 36 (62.1) | 40 (90.9) |  | 123 (98.4) | 66 (85.7) | 26 (46.4) | 37 (90.2) |  |
| **Opinion of your colleagues** | N=74 | N=20 | N=40 | N=24 |  | N=86 | N=50 | N=34 | N=23 |  |
| Has not influenced your opinion about PO yet, n (%) | 4 (5.4) | 15 (75.0) | 9 (22.5) | 3 (12.5) |  | 0 (0.0) | 0 (0.0) | 4 (11.8) | 0 (0.0) |  |
| Negative influence on your opinion about PO, n (%) | 2 (2.7) | 0 (0.0) | 1 (2.5) | 0 (0.0) | 1.31e-08 | 0 (0.0) | 0 (0.0) | 0 (0.0) | 0 (0.0) | 0.00026 |
| Positive influence on your opinion about PO, n (%) | 68 (91.9) | 5 (25.0) | 30 (75.0) | 21 (87.5) |  | 86 (100.0) | 50 (100.0) | 30 (88.2) | 23 (100.0) |  |
| **The health facility manager's recommendations** | N=105 | N=27 | N=46 | N=48 |  | N=109 | N=64 | N=38 | N=39 |  |
| Has not influenced your opinion about PO yet, n (%) | 15 (14.3) | 14 (51.9) | 7 (15.2) | 2 (4.2) |  | 2 (1.8) | 0 (0.0) | 2 (5.3) | 0 (0.0) |  |
| Negative influence on your opinion about PO, n (%) | 0 (0.0) | 0 (0.0) | 2 (4.4) | 1 (2.1) | 4.77e-08 | 0 (0.0) | 0 (0.0) | 1 (2.6) | 0 (0.0) | 0.101 |
| Positive influence on your opinion about PO, n (%) | 90 (85.7) | 13 (48.1) | 37 (80.4) | 45 (93.7) |  | 107 (98.2) | 64 (100.0) | 35 (92.1) | 39 (100.0) |  |
| **The reaction of children and/or parents/caregivers** | N=104 | N=4 | N=36 | N=20 |  | N=85 | N=20 | N=33 | N=13 |  |
| Has not influenced your opinion about PO yet, n (%) | 16 (15.3) | 3 (75.0) | 5 (13.9) | 1 (5.0) |  | 1 (1.1) | 0 (0.0) | 9 (27.3) | 0 (0.0) |  |
| Negative influence on your opinion about PO, n (%) | 35 (33.7) | 0 (0.0) | 10 (27.8) | 0 (0.0) | 0.00183 | 19 (22.4) | 2 (10.0) | 6 (18.2) | 2 (15.4) | 2.2e-05 |
| Positive influence on your opinion about PO, n (%) | 53 (51.0) | 1 (25.0) | 21 (58.3) | 19 (95.0) |  | 65 (76.5) | 18 (90.0) | 18 (54.5) | 11 (84.6) |  |

**Appendix 6.** Key elements related to qualitative CF’ acceptability.

| **Dimensions** | **Key elements** |
| --- | --- |
| Compatibility | • The tool does not conflict with existing values or customs (non-invasive tool)<br>• It complements rather than replaces the existing system<br>• Overall appreciation of new health technologies |
| Perceived complexity | • CF have little knowledge and information about the device and the project<br>• Little information is provided by HCW, and patients do not ask questions, so few know the device's purpose |
| Perceived advantages | • Improved care due to better diagnosis<br>• Avoids unnecessary trips to the PHC, and enables immediate determination of the need for hospitalization<br>• Many believe the device can diagnose or treat all illnesses<br>• Other project activities (free medications, child follow-up, etc.) are also perceived positively |
| Perceived disadvantages | • Increased consultation time (a few respondents noted decreased time): some do not appreciate this increase because they have other tasks to perform outside the center, while others see it as a sign of improved diagnosis<br>• Difficulties in financing and organizing referrals to the hospital |
| Personal emotions | • Overall confidence in the device and the HCW' ability to use it (except for some concerns when HCW had to try several times to measure saturation)<br>• Joy related to improved care for their child |
| Social influence | • High confidence in HCW, considered experts, so CF' opinions depend on them regarding health matters |

